# Point-of-care testing in a high-income country paediatric emergency department – a qualitative study in Sweden

**DOI:** 10.1101/2021.06.20.21259197

**Authors:** Reza Rasti, Johanna Brännström, Andreas Mårtensson, Ingela Zenk, Jesper Gantelius, Giulia Gaudenzi, Helle M. Alvesson, Tobias Alfvén

## Abstract

**Objectives:** In many resource-limited health systems, point-of-care tests (POCTs) are the only means for clinical patient sample analyses. However, the speed and simplicity of POCTs also makes their use appealing to clinicians in high-income countries (HICs), despite their having greater laboratory accessibility. Although also part of the clinical routine in HICs, the utility of POCTs is relatively unknown in such settings as compared to others.

In a Swedish paediatric emergency department (PED) where POCT use is routine, we aimed to characterize health care providers’ perspectives on the clinical utility of POCTs and explore their implementation in the local setting; to compare such experiences to those reported in other settings; and finally, to gather requests for ideal novel POCTs.

**Design:** Qualitative study using focus group discussions. A data-driven content analysis approach was used for analysis.

**Setting:** The PED of a secondary paediatric hospital in Stockholm, Sweden.

**Participants:** Twenty-four health care providers clinically active at the PED were enrolled in six focus groups.

**Results:** A range of POCTs was routinely used. The emerging theme *Utility of POCTs is double-edged* illustrated the perceived utility of POCTs. While POCT services were considered to have clinical and social value, the local testing practice was named to distract from the care for patients. Requests were made for novel POCTs and their implementation.

**Conclusion:** Despite their clinical integration, deficient implementation routines limit the benefits of POCT services to this well-resourced paediatric clinic. As such deficiencies are shared with other settings, it is suggested that some characteristics of POCTs and of their utility are less related to resource level and more to policy deficiency. To address this, we propose the appointment of skilled laboratory personnel as ambassadors to hospital clinics offering POCT services, to ensure higher utility of such services.

**ARTICLE SUMMARY:** *Strengths and limitations of this study:*

- This is the first study to present the use and utility of POCTs in a HIC paediatric hospital setting, directly from the perspective of its clinical staff.
- Our main finding is that deficient implementation routines limit benefit of POCT services in this well-resourced setting, strongly indicating that such deficiencies are irrespective of resource level, and more related to policy deficiency.
- The findings of the relatively small study size may be contextual, yet we believe our main conclusions to be generalizable as most of our findings are compatible with those reported in other settings.
- The study is strengthened by the diversity of its participants, corresponding to the multi-professional staffing of the PED.

## Introduction

Laboratory analyses of specific biomarkers or detection of microorganisms are a central part of clinical patient management.[1] The increasing availability of clinically graded point-of-care tests (POCTs) has brought the means for sample analyses to the point of patient care, from having previously been confined to traditional laboratory settings.[2] POCTs can be described as diagnostic tests performed near the patient or treatment facility, with a short time-to-result, that may lead to a rapid change in patient management.[3]

In many resource-limited health systems with scarce laboratory resources, POCTs for infectious diseases such as malaria, HIV, and syphilis are the only means for sample analyses.[4] Experiences with and challenges of POCTs from a health care provider perspective have been previously reported for low-and middle-income countries (LMICs), and at primary or adult care facilities in high-income countries (HICs).[5-10] However, there are, to our knowledge, no published reports of those aspects of POCTs for paediatric emergency care facilities in HICs, despite such tests being part of routine clinical practice. In such settings, POCT merits, such as short time-to-result and simplicity, have resulted in their use, despite high accessibility to advanced laboratory facilities.

Recently, we reported the use of POCTs from the perspective of health care providers in a low-income country, i.e., Uganda.[5] There, we identified several strengths and shortcomings of available POCTs and in the way they had been implemented locally. Despite some of these findings being potentially transferable to HICs, aspects of the use and utility of POCTs in HIC paediatric hospital settings remain only partially understood.

Thus, the literature has left several questions unanswered. These include: How is the routine of point-of-care testing in a HIC paediatric emergency department (PED) setting perceived by users? How do testing experiences compare to those reported in other settings? Are there any conclusions to be drawn about how POCTs have been implemented in such a setting? What requests are there for novel POCTs?

By gathering experiences and perspectives of the use of POCTs directly from end-users in a well-resourced PED setting, this study aimed to provide a puzzle piece that promotes continued discussion on how to strengthen the role and utility of POCTs in paediatric care. Ultimately, we hope our findings can contribute to an increased utility and benefit of POCTs for health care providers and care seekers alike.

## METHODS

### Setting

Stockholm is the capital of Sweden, a HIC of approximately 10.4 million inhabitants.[11] The study was conducted in 2017 at a secondary paediatric hospital housing one of three PEDs serving the greater Stockholm County, with a total population of approximately 2.4 million.[11] Along with the PED, the hospital comprises two general paediatric in-patient wards (25 beds in total), two neonatal intensive care units with two neonatal wards, several in-house paediatric out-patient units, and three additional satellite out-patient units.

The PED is visited by 30,000 children aged 0-18 years annually, infections being the leading cause for consultation. It is staffed by paediatric consultants/specialists, residents, newly graduated junior medical doctors (filling a temporary position to qualify for clinical rotations leading to medical license), nurses, and nurse assistants. Outside office hours, the PED is staffed by nurses, nurse assistants, junior doctors, and one to two residents/specialists managing the PED as well as the in-patient wards, the neonatal units, and the adjacent delivery and antenatal care units of the hospital. During these hours, paediatric and neonatal consultants are on-call, ready to support the physicians on site by telephone. In addition to centralized laboratory and radiology functions being present at the hospital, the PED and wards at the hospital are equipped with a variety of POCTs.

### Study design and participants

All health care providers, including nurses, nurse assistants, senior paediatricians (consultants), junior doctors, and paediatric residents, clinically active at the PED were invited to participate in this study through purposive sampling, by e-mail, or via invitations at staff meetings. In total, 24 individuals agreed to participate and were placed into one of six focus groups, according to profession, to promote professional homogeneity.[12, 13] The groups were composed of three to six participants (Table 1) and engaged in moderated qualitative focus group discussions (FGDs) in Swedish from March to December 2017. Five of the FGDs were held in a conference room at the hospital, and one FGD (with consultants) was conducted off-site during an annual clinic gathering.

**Table 1.**
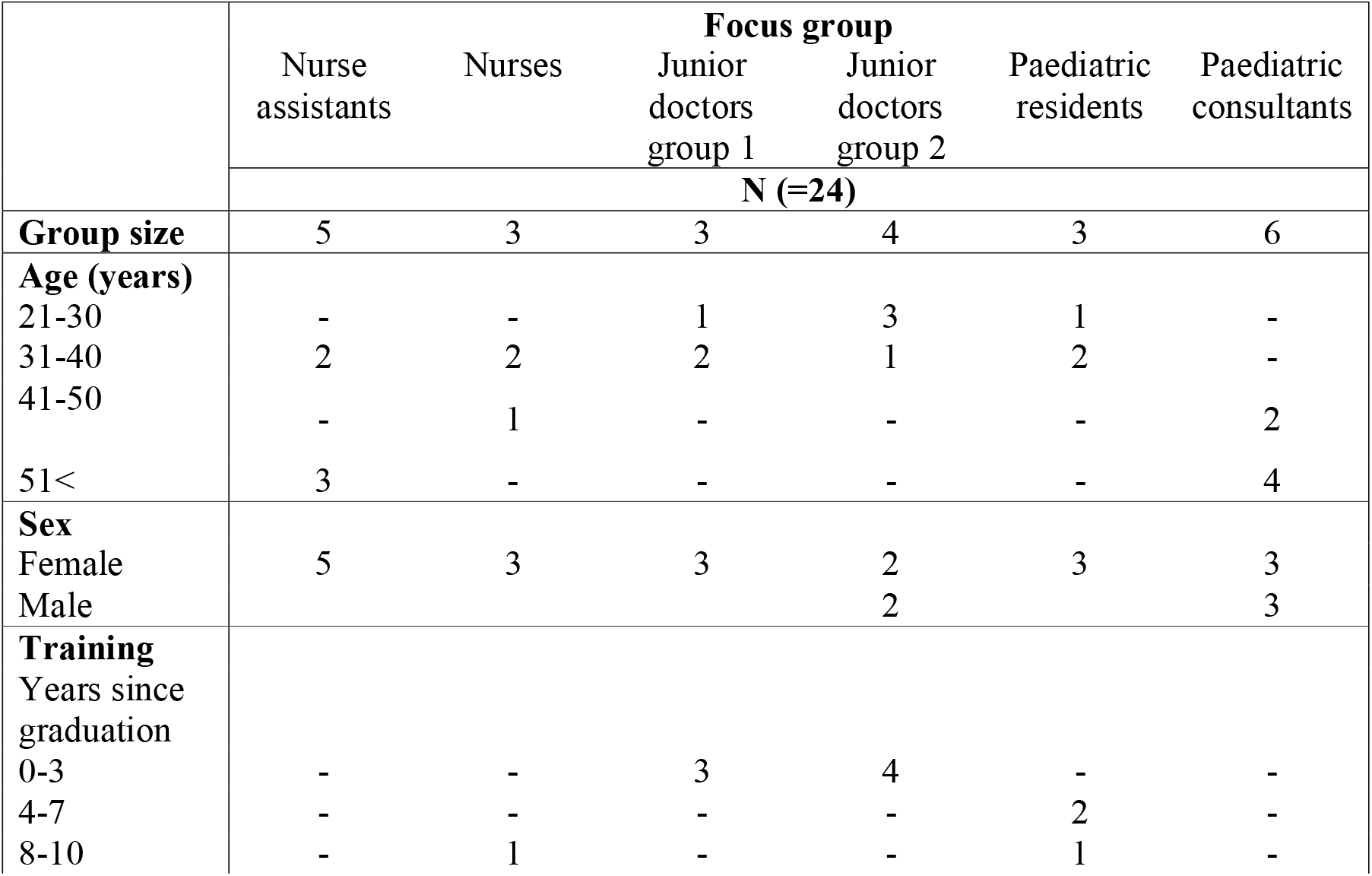
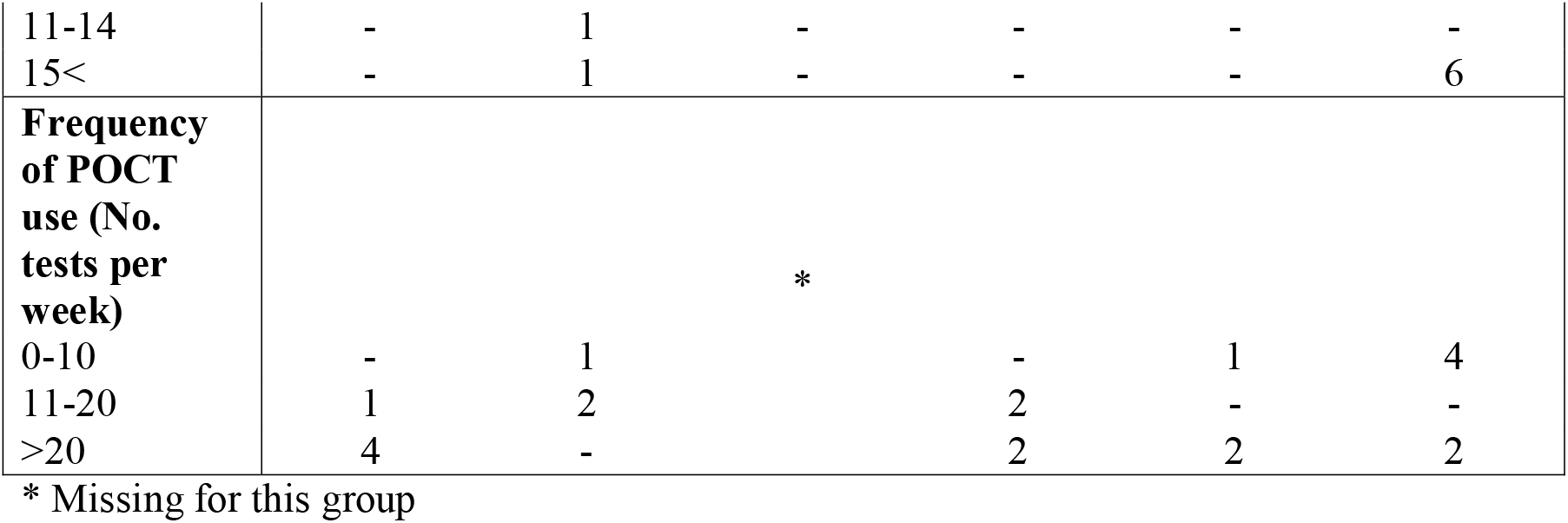
Composition of the focus groups and characteristics of the participants.

### Data collection and analysis

Based on reports of studies with similar approaches and following discussions within the study team, an interview guide (Appendix 1) was developed, with the main topics being: *experiences of using current point-of-care technology; what is most important when point-of-care tests are used; what an ideal point-of-care test would include*. The interview guide was pilot tested in the ‘Junior Doctors group 1’, before being used in the subsequent FGDs.[5, 7, 14, 15] All FGDs were moderated by either two or three of the authors (RR, HMA, JB), with one moderator taking an active role and the other(s) having supporting roles. Initiating each discussion, moderators presented the purpose of the study and the general forms for discussions, and defined POCTs according to Schito et al.[3] The FGDs were audio-recorded (median duration 74 minutes, range 60 to 81 minutes) and transcribed verbatim in Swedish by RR and JB.[12, 16, 17]

An inductive data-driven approach for content analysis was taken for coding transcripts, using NVivo for Mac v. 11.4.3 (QSR International) software.[18, 19] Meaning units in Swedish were identified and coded in English. Once all data had been coded, each code was re-evaluated and compared to the others. This process was repeated several times by RR upon discussions with HMA, resulting in the fusion of matching codes and re-coding of meaning units whose substances were better understood after repeated reading. The obtained set of codes was then abstracted into subcategories, categories, and an overarching theme. Following continued discussions within the study team, the abstraction was revisited on several occasions until agreement of saturation had been reached.[18]

### Ethics

The study was approved by the Regional Ethical Review Board in Stockholm (ref. 2016/2296-31/1). All participants provided written informed consent and were compensated with four movie vouchers each. Soft drinks and pastries were served during FGDs. All transcripts were de-identified during data transcription and in the resulting manuscript.

### Patient and public involvement

Patients or members of the public were not involved in this study.

## RESULTS

Data analyses yielded nine subcategories that were grouped into two categories and abstracted into one theme: ‘*Utility of POCTs is double-edged*’ (Table 2). Categories and subcategories are described in detail below. In addition to the these, contextual information describing the availability of POCTs, and in how they are taught to and used by the participating professions are illustrated in sections: ‘*POCTs available and unavailable to us*; and ‘*How we are taught about POCTs and who among us do the testing’*. Requests for novel POCTs and for their use are presented in ‘*Requests for POCTs’*.

**Table 2.** Structure of theme, categories, and subcategories

| **Table 2. Structure of theme, categories, and subcategories** |  |  |
| --- | --- | --- |
| **Theme** | **Main categories** | **Subcategories** |
| Utility of POCTs is double-edged | POCTs have clinical and social values | Paediatric care is favoured by the use of POCTs |
|  |  | Fulfilling personal needs of doctors |
|  |  | Reassuring guardians and fulfilling their expectations |
|  | Our current testing practice distracts from care for our patients | The temptation of POCTs disrupts our clinical reasoning |
|  |  | POCT results cannot always be trusted |
|  |  | Non-standardized use of POCTs |
|  |  | POCTs are not “all good” |
|  |  | We are inadequately informed about our POCTs |

### POCTs available and unavailable to us

In total, 18 different testing procedures were identified by participants as POCTs and available to them, with additional eight tests being known from other Swedish workplaces (Table 3).

**Table 3.** POCTs mentioned in at least one FGD as available to participants and as known from other settings.

|  |  |
| --- | --- |
| **POCTs available to participants** | **POCTs known from other Swedish clinics** |
| C-reactive protein (CRP) | Leucocytes |
| Blood gas | Chlamydia trachomatis |
| Urinalysis | Urine drug screening |
| mariPOC® Respi test | Full blood count |
| Respiratory Syncytial Virus (RSV) | Wider blood gas panel |
| Pregnancy test | Haemoglobin |
| Glucometer | Erythrocyte sedimentation rate |
| Influenza | Transcutaneous carbon dioxide |
| Urine drug screening |  |
| Streptococcus group A |  |
| Haemoglobin in faeces |  |
| Blood ketones |  |
| Malaria |  |
| Mononucleosis spot |  |
| Transcutaneous bilirubinometer |  |
| Haemoglobin in blood |  |
| Uricult test |  |
| Saturation meter |  |

### How we are taught about POCTs and which of us do the testing

There was not a given uniform method for learning about POCTs and being trained on how to use them. Learning along the way was the most frequently named learning method:

> *It is like a continuing habituation to a new test, when you start using it and then you incorporate it into your arsenal of things to use in diagnostics*.
>
> (Consultant group)

Furthermore, junior doctors were described as learning from senior colleagues or nurses, and nurses and nurse assistants from peers or through a hospital online teaching platform.

Regarding the testing procedure, most sample acquisitions (besides throat swabbing, which some of the doctors performed) and test analyses were described as being conducted by nurses, or by nurse assistants through special delegation from nurses.

### POCTs have clinical and social values

Participants described POCT characteristics and use as favourable to meeting clinical needs in paediatric settings, as well as serving non-clinical interests, fulfilling the needs of care seekers, and fulfilling the personal needs of doctors.

#### Paediatric care is favoured by POCT use

POCTs were described as easy to learn and use, and as beneficial to paediatric clientele. The latter was due to POCTs being less invasive (e.g., capillary blood samples, or transcutaneous bilirubinometer) and requiring smaller specimen volumes than those required by the central laboratory.

While reflecting on the influence of POCTs on patient management, participants credited them with clearing up ambiguous situations and facilitating patient assessment.

> *It is also such that babies, or babies and children in general, can be somewhat difficult to interpret sometimes. … E*.*g*., *they can say that they have a stomachache, but that could be anything! And then it is very pleasant to have the dipstick* [urinalysis POCT] *just to rule out a urinary tract infection. I think many of these tests are for ruling out more serious…*
>
> (Junior doctor group 2)

Furthermore, POCTs were described as reliable, rapid, and, in some cases, multiplex. Participants described them as simplifiers and accelerators of differential diagnostics and as guides to proper management and treatment of patients. Also, their quickness was described as shortening patient time spent in the PED. Specific situations where POCTs were described as especially useful were in triage and emergency situations. The CRP test was indicated to be an aid in fever case management and for evaluating treatment response in patients admitted due to infection, and the blood gas test for following the progress of patients in respiratory distress.

#### Reassuring guardians and fulfilling their expectations

All focus groups witnessed patient guardians specifically requesting diagnostic tests to be undertaken, during consultations. It was perceived that some guardians consider testing to be an essential part of patient assessment. It was also suggested that some guardians bring their children to the PED only to have them undergo testing, and several participants described decisions on testing sometimes being based on such requests. At the same time, test results were considered to have a reassuring effect on worried guardians and a pedagogical role in communication between caregivers and care seekers.

> *Sometimes I think it can be purely communicative with guardians. It can be that we now have taken an infection test, and it is low; this very much looks like a viral infection combined with having had symptoms for a couple of days, and it is low, so it doesn’t suggest bacterial infection. You can go home and rest and come back if there were something*.
>
> (Resident group)

#### Fulfilling personal needs of doctors

In addition to being used for the benefit of patient management and fulfilling care seeker needs, POCTs were also viewed as having personal value to doctors, especially to those with little clinical experience. The credibility of assessments and clinical decisions made by junior doctors was described as being strengthened by test results.

> *The technology is hard to beat, you can say. Either that or grey hair; I’m waiting for either one*.
>
> (Junior doctor group 2)
>
> *Let’s say that you are at home, and are called by a very inexperienced, new colleague; then I think you get like, ‘but maybe we should take some extra samples’, because I’m not really sure of the anamnesis you get, because I don’t really know, I don’t know that person as much as if I had with a more experienced colleague where I would have trusted the story more and everything. I can imagine that it gets like that*.
>
> (Consultant group)

Elaborating on testing for the benefit of the doctor, test results were thought to reduce doctors’ anxiety regarding mistakes, and as having an educational function, especially for less experienced doctors who were considered most prone to prescribing tests.

> *So, I feel that sometimes you hope to be allowed to take some extra test just to receive that feedback for yourself. But that it is mostly for me, like for my learning. For the future*.
>
> (Junior doctor group 1)

### Our current testing practice distracts from our care for patients

Concurrent with observing the benefits of POCTs for work at the paediatric clinic, participants expressed concerns regarding their use, and how POCTs could complicate things. The subcategories of this topic are illustrated by a sample of quotes in Table 4.

**Table 4.**
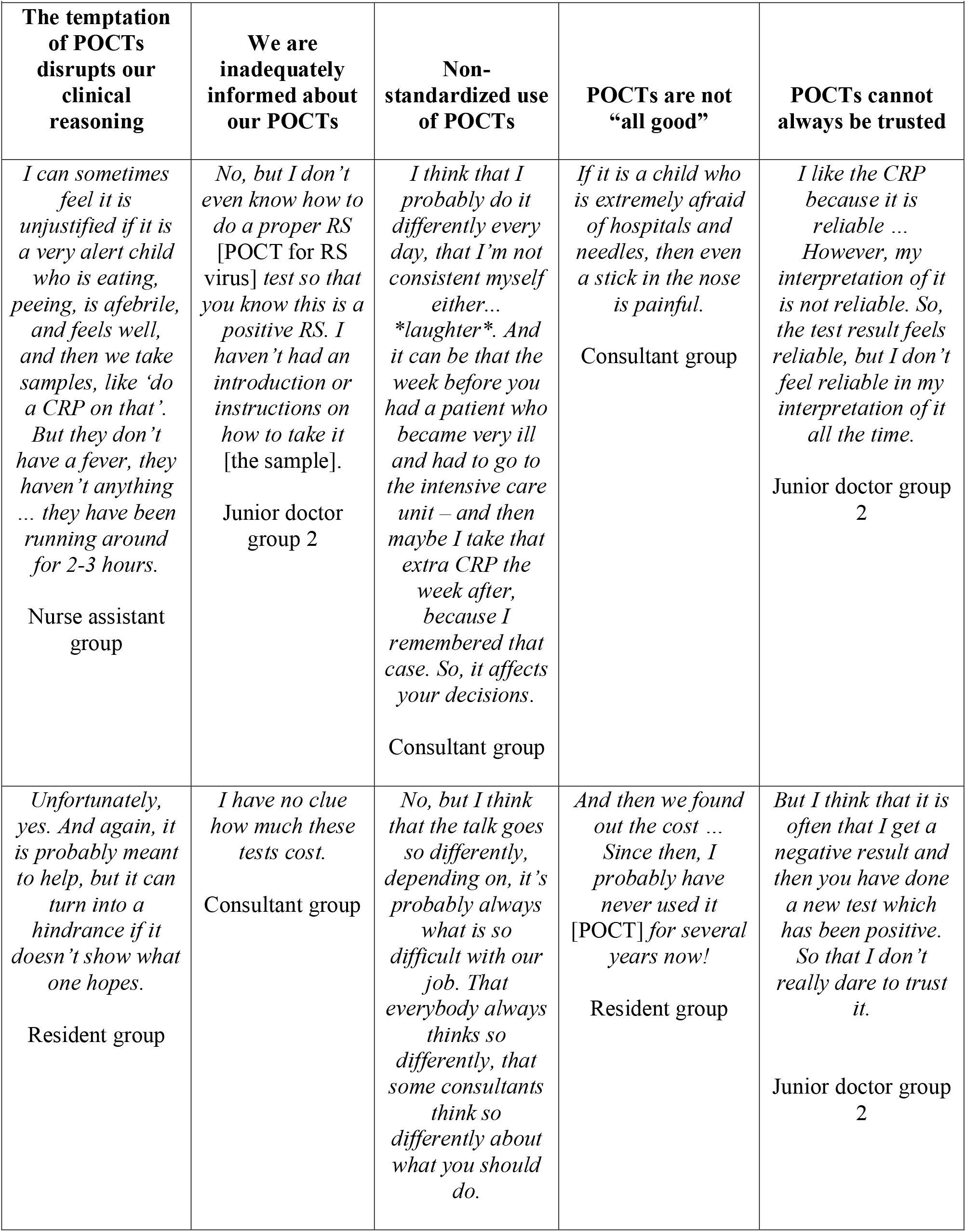
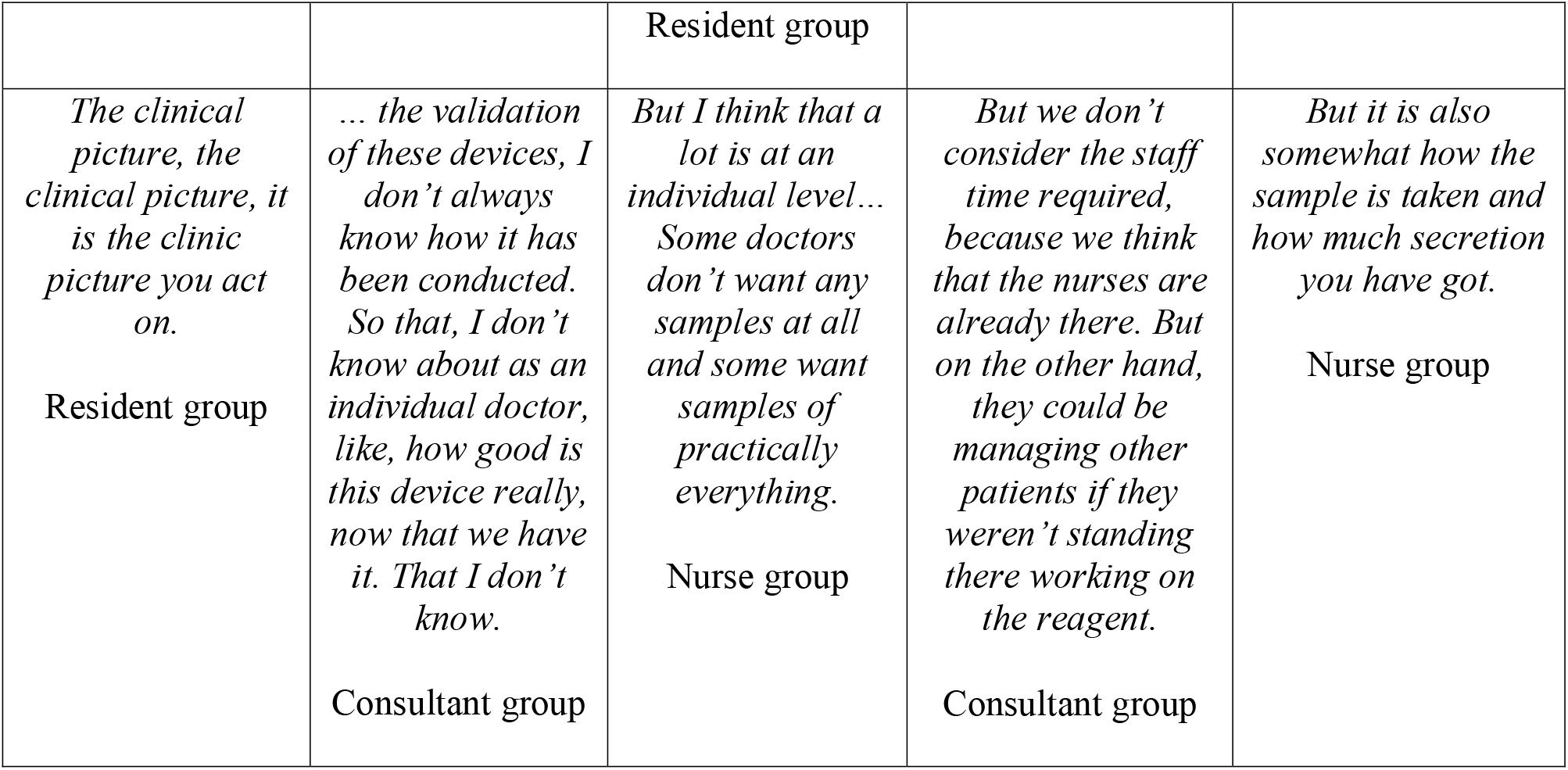
The subcategories of ‘Our current testing practice distracts our care for our patient’ with a sample of corresponding quotes.

#### The temptation of POCTs disrupts our clinical reasoning

The availability, rapidness, and simplicity of POCTs were described as allowing for overuse and unjustified testing, rather than relying on clinical skills. Such practices were described as a burden to nurses and nurse assistants, and subjected children to unnecessary procedures.

Concurrently, test results were described as less decisive for patient management than what could be observed in the clinical picture.

Contrary to descriptions of POCTs clearing up ambiguous situations, they were also viewed as sometimes resulting in increased uncertainty. In some instances, wrongful testing procedures caused incorrect results that resulted in poor decisions. In other cases, results differed from what was expected, leading to doubt of clinical assessments and instigating further investigation. These concerns were in line with others relating to the difficulty of interpreting test results and deciding on proper action.

#### We are inadequately informed about our POCTs

Participants recognized personal knowledge gaps concerning how POCTs work, their accuracy and cost, and the range of tests available to them. Some also stated that they had not been taught the correct sampling techniques or analysis procedures (Table 4).

#### Non-standardized use of POCTs

Participants with experience at other workplaces described their current paediatric setting as being more reliant than adult clinics on POCT analyses, and of differing testing routines between paediatric hospitals. They also described a lack of written clinical guidelines for when and why to use POCTs. This was reflected by testimonies of in- and inter-person non-uniformity among doctors regarding using POCTs. As junior doctors had been described more likely to use POCTs, there were also descriptions of non-stringency in patient management by senior doctors (Table 4). Also, and in the absence of written guidelines, junior doctors were said to receive different instructions depending on which senior colleague they had consulted. Another more philosophical explanation given for the lack of uniformity was the practice of medicine as an art.

#### POCTs are not “all good”

Contrary to prior descriptions of favourable characteristics of POCTs, participants also described the tests as expensive, resource-intensive, and uncomfortable for children. Cost was also stated as an inhibitor of test utility.

#### POCT results cannot always be trusted

Addressing accuracy, POCTs were described as quicker, but their results less trustworthy than those of laboratory analyses. Also, sometimes POCT results were considered difficult to read, allowing for misinterpretation. POCTs identified as having low accuracy were those for RSV, urinalysis, Streptococcus group A, and Mononucleosis spot.

### Requests for POCTs

At the end of each discussion, participants were asked to name existing or non-existing POCTs that they would like to be made available to them, as well as to state ideal features of POCTs with high utility (Table 5). The possibility of POCTs becoming available to the general public and used outside clinical settings was briefly addressed by the ‘Consultant group’. This raised concern that it would burden health care providers with worried persons seeking care due to test results that they were not qualified to interpret. However, POCTs for self-use, when used in conjunction with video consultations by health care providers, were thought to have a role in the not-too-distant future, and that such a scenario could also benefit patients in low-income countries with lower access to health services. Relating to requested strategies for using POCTs, participants called for clinical protocols for their use, emphasizing the need for value of patient management for each conducted test.

**Table 5.** Specific POCTs and their characteristics, as requested by participants

| ***Table 5. Specific POCTs and their characteristics, as requested by participants*** |
| --- |
| **Specific POCTs requested** |
| Scanner of airways<br>Point-of-care ultrasonography<br>Appendicitis POCT<br>Cancer POCT<br>Migraine POCT<br>Procalcitonin POCT<br>Blood POCT for meningitis<br>A more rapid intoxication POCT<br>POCT for creatinine<br>Rapid PCR POCT for urinary tract infections (UTI)<br>Infection aetiology POCT<br>Skeletal fluoroscopy for fracture detection<br>Transcutaneous UTI scanner of urine bladder |
| **Desired features of ideal POCTs** |
| Multiplex<br>Automated analyses<br>Low-cost<br>Non-invasive<br>Quick<br>Highly accurate<br>Fool-proof<br>Comfortable for patients<br>Should direct towards correct management<br>Should quicken diagnostics<br>Test results should be automatically entered into electronic medical record of patients<br>A treatment-guiding POCT |

## DISCUSSION

This is, to our knowledge, the first study illustrating how POCTs are perceived by end-users in a high-resource paediatric emergency hospital setting. Here, the praxis of POCT-driven diagnostics is a normalized part of daily operations, with a range of different tests in use. Although part of the routine clinical practice, our participants perceived the utility of POCTs as double-edged: on one hand, being beneficial to patient management in paediatric emergency care, and having reassuring value to health care providers and care seekers; on the other hand, being a distraction in the work at hand, with little stringency in when and why POCTs are used. Furthermore, we illustrate a value of diagnostic testing that is not strictly clinical. Finally, we give recommendations for increased quality of POCT services and present requests for ideal POCTs and their use.

Despite POCTs being considered especially beneficial to resource-limited health systems of LMICs, this study, in line with others, shows their use to be appreciated also in a well-resourced context, despite high accessibility to more advanced laboratory diagnostics and skilled personnel.[4, 5, 7, 8, 20-23] POCTs are often less invasive and require smaller volumes of patient specimens than central laboratory facilities, which, together with their evolving multiplexity, promotes their use in paediatric clinics.[2, 24] In our study, POCTs were merited as facilitators of patient management by accelerating differential diagnostics and patient flow at the PED, being favourable to paediatric clientele and essential to the emergency department setting of this study. Such merits have previously been described in primary care.[9]

Meanwhile, concerns were raised regarding the accuracy of POCT results, challenges in their read-outs, insecurities in their proper use, and unexpected test results. Such concerns, together with descriptions of how POCTs can increase clinician uncertainty and cause unnecessary ancillary investigations or incorrect assessments, are compatible with prior reports.[5, 10, 25, 26] Concurrently, we found knowledge gaps among our clinicians regarding essential aspects of the POCTs that they are using daily. These included insights into test accuracy and cost, awareness of correct testing procedures, and an understanding of how the analyses are performed by the assays. Unexpectedly, these knowledge gaps seem to be irrespective of clinician seniority. Despite the benefit of POCTs not requiring advanced laboratory skills, it is evident that the fundaments of POCTs have not been introduced to end-users, and that teaching forums also need to be conducted for clinicians, in order to enhance the quality of POCT services.[2]

Our findings also highlight a lack of stringency in the prescription of POCTs; we believe this to be partly explained by inadequate training routines of end-users as well as the absence of testing protocols. Furthermore, the absence of such protocols is considered to allow for what one participant described as practicing the “art of medicine”. As such a practice permits incoherencies in the management of patients and can be viewed as posing a risk to patients, one might also argue that clinical medicine is seldom straightforward, and that there will always be differences in its practice.[27]

As the described deficiencies in currently available POCTs and in their use have been reported in low-, middle-, and high-income countries,[5-10, 25, 26, 28] it can be concluded that there are flaws in the design of currently available POCTs and their utilization that are universal and irrespective of available resources. Also, it is evident that insufficient implementation processes are not solely due to strained resources, but rather that they have not received adequate attention by stakeholders. This could have negative consequences for the adoption of new technologies and creates barriers to the full utility of such devices.[29] The use of POCTs should be viewed as part of a diagnostic *service* offered by caregivers, and hurdles to the success of such services are intertwined with the challenges faced by the health system in which they are embedded.[4]

As the accuracy of POCTs is limited by the know-how of its user, the introduction of protocols for training and certification of end-users with requirements of regular renewal of such would minimize the risk of human error, improve quality control measures, and adhere to requirements for ISO 22870 and 15189 accreditations.[2, 24, 30, 31] To ensure the quality of such measures, while building on recommendations by Larsson et al.,[2] we propose that hospital laboratory units appoint ‘ambassadors’ to clinics offering POCT services. Their duties should include an inventory of local POCT needs, the procurement and implementation (including staff training) of POCTs, and repeated quality control of the assays and in their use. Such a task needs to be complemented by the inclusion of POCTs into existing and future patient management protocols, where applicable. Being aware of the difficulties of adhering to such recommendations in LMICs, they could arguably be feasible in high-resource settings.

As Lupton contends, medical technology has a major role in health care delivery and is integral to the experiences of caregivers and care seekers alike.[32] Furthermore, Armstrong et al. illustrate how the use of diagnostic instruments can have social functions, such as fulfilling clinician duties to patients,[27] and there are numerous reports on how diagnostic testing influences care seeker satisfaction.[6, 9, 10, 24, 26, 33, 34] Such social values are also recognised by our participants, illustrating how test results can curb the insecurities of less experienced doctors, strengthen their credibility in dialogue with senior colleagues and care seekers, help them gain clinical expertise, and increase care seeker satisfaction. At the same time, clinically unjustified testing adds to the workload of the personnel and subjects children to procedures deemed invasive enough to be uncomfortable to them.[35, 36 Also, reports of over-reliance on technology as a cause of de-skilling clinicians are echoed by our findings stating that doctors would need to rely more on their clinical skills in the scenario in which POCTs are not available to them.[5, 10, 26, 37] Evidently, it is difficult to cater to clinical and social needs of testing, while avoiding the risks and disadvantages of unwarranted testing.

Interestingly, most of the features requested by our participants regarding *ideal* POCTs are consistent with requests made by Ugandan health care providers.[5] Both settings requested non-invasive, cheap, quick, foolproofly and accurate tests with the ability to direct clinicians towards proper patient management. Regarding specific conditions for which POCTs were requested, there were contextual differences between the two settings, reflective of differing epidemiology and availability of laboratory analyses.

## STRENGTHS AND LIMITATIONS

Ideally, there should be four to eight participants in each focus group.[12] Although additional participants had repeatedly been invited to the FGDs, there were last minute absentees and other obstacles to enrolling more participants, mainly due to the irregular working hours of our target participants. Yet, we consider 24 to be a large enough sample, and even though larger groups could have produced additional perspectives, they could also have limited the depth of discussions. Furthermore, as we only investigated one hospital, some of our findings could be contextual. However, as most of our findings are compatible with those from other settings, we believe our main conclusions to be generalizable, despite the possibility of differing testing practices in other paediatric hospitals.

The study is strengthened by the diversity of its participants, corresponding to the multi-professional staffing of the PED, and their grouping according to profession, promoting participants to speak freely. Since authors RR (paediatric resident) and JB (medical student and employed as nurse assistant) were employed at the hospital at the time of this study, their familiarity with the participants further promoted open and friendly discussions. As author HMA was previously unknown to participants, her presence, qualitative experience, and non-clinical profession (medical anthropologist) helped keep discussions on track and limited jargon.

## CONCLUSION

In a Swedish paediatric emergency department setting, a range of POCTs is routinely used in clinical practice. While the utility of POCTs is seen as double-edged here, it is shown to have clinical and social value. However, deficient implementation routines limit the benefit of POCT services. As most of our findings are shared with LMICs, it is suggested that some characteristics of POCTs and of their utility are less related to resource level and more to policy deficiency. To address this, we propose the appointment of skilled laboratory personnel as ambassadors to hospital clinics offering POCT services, to ensure higher utility of such services.

## Supporting information

SRQR checklist

## Data Availability

De-identified transcripts in Swedish of audio files will be provided upon request to the corresponding author. Audio files will not be provided as they contain identification of participants.

## ACKNOWLEDGMENTS

The authors thank all participants of the study, and to the TREND (Trial of Respiratory infections in children for ENhanced Diagnostics) study group. Our gratitude also goes to Region Stockholm (combined residency and PhD training to RR, and ALF grant 20150503), whose financial contributions made this study possible, and to Lamont Antieau for language editing.

## AUTHOR CONTRIBUTIONS

RR, JB, HMA and TA conceptualized and designed the study with input from AM, IZ, and JG. The interview guide was developed by the abovementioned. RR, JB and HMA moderated focus group discussions and conducted data collection. RR and JB transcribed audio files and managed the data. RR analysed and interpreted the data upon discussions with HMA. The analysis results were revisited and repeatedly discussed with all authors before being finalized. RR wrote the manuscript with input from all authors. The entire team of authors reviewed and approved the final manuscript.

## COMPETING INTERESTS

None declared.

## FUNDING

Region Stockholm (grant numbers K0175-2016 and ALF-20150303).

## APPENDIX 1

Final interview guide used during focus group discussions.

### Experience using current point-of-care technology

A. *What point-of-care tests do you use today? At the emergency department? In the wards?*
B. *What is your experience with these tests? What aspects do you value? What aspects do you not like?*
  a. *Consider how confident you are in the POCTs: which are the tests you are most confident in using? Least confident? How is reliability? Which POCT provides the most accurate answer and which does not?*
C. *How did you learn to use POCTs? How was the process of learning? Was it easy to learn, and how long did it take? Who taught you how to handle the tests?*
D. *Are there currently available POCTs that you are aware of but do not use? Which? And if so, why?*
E. *Are there any guidelines/protocols for using POCTs? Have they changed over time?*

### What is most important when you use POCTs?

A. *What are the most important characteristics of a POCT to you?*
  a. *What do you consider the most undesirable characteristics of a POCT?*
B. *Which aspects of POCTs, or concerns about them, makes it less likely for you to use them?*
  a. *Difficult to manage, unpleasant for patients, expensive, or other…?*
C. *In your experience, are there any concerns in health workers when using POCTs? Which?*
  a. *Are there any POCTs you prefer to take twice? Or complement with other examinations?*
  b. *PROBE: What defines “unnecessary sampling”? How do one know it is unnecessary?*
D. *Which are the most important clinical decisions that POCTs help you make?*
E. *In which situations in clinical practice do you believe POCTs are most helpful?*
  a. *If there was no POCTs, what difference would it make at the clinic?*
  b. *How does having POCTs available affect your other duties/your workload?*
  c. *PROBE (for senior doctors): Does the need of POCTs differ when managing a patient directly in contrast to supervising a junior doctor at a distance (e*.*g*., *over the phone)?*
F. *How effective and useful do you think your colleagues believe the POCTs are?*
G. *Are there differences among your colleagues in what they think about POCTs?*
  a. *If you compare junior doctors to seniors – have you noticed differences?*
  b. *Or within different specialties?*
  c. *Emergency department compared to wards?*
H. *For whom are POCTs performed mostly? (For diagnostics, for the patient, for the guardians, for the doctor, or for the senior colleague?)*
I. *Who can order a POCT, carry out the test, communicate result to care seeker? What is your opinion of this division of labour?*
J. *Communicating with the patient/care seeker – does the use of POCTs affect your relationship? Do care seekers ask about your choice of tests? Ask for it?*
  a. *Do you explain about what you do to the care seeker?*
  b. *Do POCTs affect the amount of time you have with your patients?*
  c. *Which examinations are replacing the POCTs? Who would perform them?*
  d. *POCTs as pedagogical tools, who uses them?*

### The ideal POCT

A. *How would an ideal POCT improve clinical practice?*
B. *Which conditions or diseases would you like an ideal POCT to diagnose?*
C. *Which characteristics would be desirable in ideal POCTs?*

### Any other questions?

## Notes

### Competing Interest Statement

The authors have declared no competing interest.

### Author Declarations

Regional Ethical Review Board in Stockholm (ref. 2016/2296-31/1).

